# Quantitative, multiplexed, targeted proteomics for ascertaining variant specific SARS-CoV-2 antibody response

**DOI:** 10.1101/2022.02.14.22270845

**Authors:** Ivan Doykov, Justyna Spiewak, Kimberly C. Gilmour, Joseph M Gibbons, Corinna Pade, Áine McKnight, Mahdad Noursadeghi, Mala K Maini, Charlotte Manisty, Thomas Treibel, Gabriella Captur, Marianna Fontana, Rosemary J Boyton, Daniel M Altmann, Tim Brooks, Amanda Semper, James C Moon, Kevin Mills, Wendy Heywood

## Abstract

Determining the protection an individual has to SARS-CoV-2 variants of concern (VoC) will be crucial for future immune surveillance and understanding the changing immune response. As further variants emerge, current serology tests are becoming less effective in reflecting neutralising capability of the immune system. A better measure of an evolving antigen-antibody immune response is needed. We describe a multiplexed, baited, targeted-proteomic assay for direct detection of multiple proteins in the SARS-CoV-2 anti-spike antibody immunocomplex. This enables a more sophisticated and informative characterisation of the antibody response to vaccination and infection against VoC. Using this assay, we detail different and specific responses to each variant by measuring several antibody classes, isotypes and associated complement binding. Furthermore, we describe how these proteins change using serum from individuals collected after infection, first and second dose vaccination. We show complete IgG1 test concordance with gold standard ELISA (r>0.8) and live virus neutralisation against Wuhan Hu-1, Alpha B.1.1.7, Beta B.1.351, and Delta B.1.617.1 variants (r>0.79). We also describe a wide degree of heterogeneity in the immunocomplex of individuals and a greater IgA response in those patients who had a previous infection. Significantly, our test points to an important role the complement system may play particularly against VoC. Where we observe altered Complement C1q association to the Delta VoC response and a stronger overall association with neutralising antibodies than IgG1. A detailed understanding of an individual’s antibody response could benefit public health immunosurveillance, vaccine design and inform vaccination dosing using a personalised medicine approach.

## Introduction

After the first cases of SARS-CoV-2 were identified in late 2019, 2020-2021 saw the development and roll out of the world’s fastest and largest global vaccination programs. However, with potential waning immunity over time (*1*) and the impact of infection from emerging Variants of Concern (VoCs) (*2*), it is apparent that there is a need for better and more informative testing (*3*). This will help determine the clinical need for booster vaccination and timing of the boost itself. First generation tests were rolled out at scale but are largely based on simple non-specific binding to the prototypic Wuhan-Hu-1 spike sequence. It is now clear that these methods overestimate the actual protective immunity against VoC (*2*). Current technologies that directly measure accepted correlates of protection such as neutralising antibodies (NAbs) scale poorly for clinical utility, while serological approaches (ELISA or ECLIA), measure only part of the antibody response and omit measurement of effector Fc antibody functions such as complement involvement (*4–6*).

To aid in understanding the antibody response to SARS-CoV2 we have developed a methodology that includes a ‘bait and capture’ system, followed by a multiplexed and targeted proteomic liquid chromatography tandem mass spectrometry (LC-MS/MS) analyses. The combination of immunocapture with the multiplexing capability to look at multiple proteins involved in the immune response and high accuracy of mass spectrometry quantitation, makes this an extremely powerful and more informative combination. In addition, tandem mass spectrometers are also routinely used for small molecule clinical assays in most UK pathology laboratories and are therefore platforms which could be utilised for targeted proteomic assays. It’s only recently, with improving technology that they are becoming recognised for their potential clinical application for multiplex protein analysis (*7*).

In this work we describe how we have used this novel assay to compare with previously determined immune correlates (*2, 8*) in serial samples, in response to vaccination, infection and an individual’s potential protection against VoC. This analysis was performed using serum samples from the COVIDsortium study (*8–12*) where previously detailed longitudinal immunological analysis had been carried out. This unique cohort included healthcare workers with and without laboratory confirmed SARS-CoV-2 infection, during the first UK wave with the Wuhan Hu-1 strain and after one and two dose vaccination (Pfizer/BioNTech BNT162b2) (*8, 10, 12, 13*). Our analyses demonstrates that the conventional measurement of IgG1 is insufficient to determine an individuals complete immuno-response or protection due to infection and vaccination. We show that responses to infection and vaccination can be heterogeneous from person to person and by broadening the portfolio of those biomarkers involved in the monitoring of the immune response, this assay provides a more informative picture of immune responses. This assay could be used to determine an individual’s immune potency against VoC but also aid in design of future vaccine trials.

## Results

### The development of a multiplex LC-MS/MS Assay for measuring antibody mediated response to SARS-CoV-2 spike antigen

The multiplex assay is significantly more sophisticated and informative because of its ability to quantitate simultaneously and accurately, all major antibody species and their subclasses, as well as key components of the downstream complement pathway. The rationale behind inclusion of complement proteins is due to their involvement in formation of antigen-antibody complexes. Figure 1A is a schematic representation of how the assay captures and analyses a patient serum immunocomplex specific to the SARS-CoV-2 spike region. A recombinant SARS-CoV-2 S1 spike protein from any VoC is first bound to a 96-well plate in a simple procedure described below. Patient serum is incubated with the bait for 60 min to capture the immunocomplex and non-specific proteins washed away. All immunocaptured proteins including the spike, are trypsin digested and the unique signature peptides analysed by a targeted LC-MS/MS analyses. The assay was capable of identifying and quantitating the immunoglobulins IgG1, 2, 3 & 4, IgA1, IgM and the complement factors C1q, C4b, and C9 using 10 μl of serum with a CV range of 1.68%-13.6% for high response QCs and 1%-15.3% for low response QCs. To improve reproducibility over existing immunodetection assays, results were expressed as a ratio of each immunological protein to the SARS-CoV-2 spike (using a S1 peptide common to all variants). This improves the CV % by a factor of ∼ 10-fold i.e. a standalone value CV for IgG1 is reduced from 13.8% to 3.4% if the value is ratioed to spike bait. Internal monitoring of bait binding also provides a quality assurance for plate preparation.

**Figure 1.**
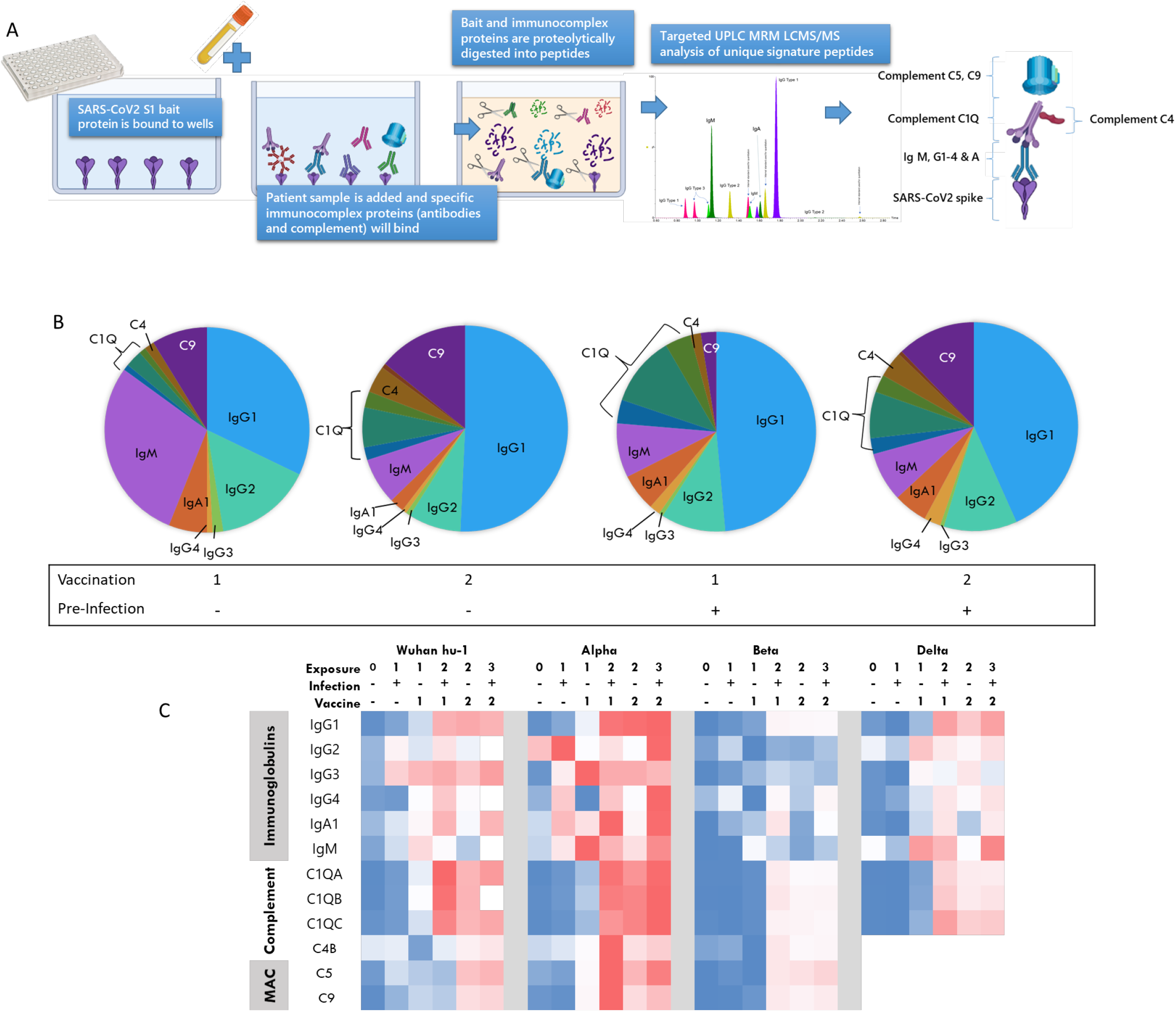
Principle of the targeted LC-MS/MS Immunocomplex assay. **(A)** shows a summarised workflow and schematical representation of the bait-capture-LCMS/MS assay. **(B)** Composition of the immunocomplex at different vaccination and pre-infection stages. Mean values used for each protein. **(C)** Heatmap of all proteins measured in the multiplex as determined by normalised mean values of the protein ratioed to spike peptide against each S1 bait variant and vaccination group MAC= membrane Attack Complex. Blue to Red colour scale indicates lowest to highest values.

Figure 1B shows how the profile of the immunocomplex changes according to vaccination and prior infection. A heatmap summary of the proteins included in the immunocomplex and their response to exposure and variants is given in Figure 1C and summarises the increase of the immunocomplex with increasing exposure against all SARS-CoV-2 variants. However, a notable reduction of detectable immune activity against the Beta VoC can be observed compared to other variants analysed. A strength of our assay is to measure all major antibody types simultaneously, allowing comparison of inter-individual and exposure isotype heterogeneity. When all components of the immunocomplex are viewed collectively (supplementary figure S1) a wide variation in those with 2 or more antigen exposures can be observed with clear outliers. Closer inspection of the outliers highlights that these individuals have an atypical immunocomplex profile with one interesting individual who has a dominant IgG4 response. IgG4 is associated with anti-inflammatory properties as it can undergo fab-arm exchange thereby limiting effector functions (*14*). This indicating that a ‘one size fits all’ strategy using the current Elecsys assays does not provide us with enough information if we are to determine and study an individual’s current immune status.

As proof of principle, we also demonstrated that this methodology can also be applied to the much less invasive measurement of antibodies and immune response proteins (than collecting blood serum) using dried blood spots, saliva or saliva adsorbed onto Guthrie card blood collection strips or “lollipops” (supplementary figure S2).

### Comparison with the gold standard S1 RBD serology assay and authentic live virus neutralisation

We compared our assay with the gold standard serology and authentic live virus neutralisation assays at three timepoints, sampling responses 8 weeks after natural infection with Wuhan Hu-1 during the first UK wave and after the first and second dose vaccination. This enabled us to evaluate and compare our assay after one to three antigen exposures using the same existing peer reviewed, published underpinning datasets analysed in these cohorts (*8, 10, 12, 13*). We compared results from our assay to those obtained by second generation serology, which measures a total response to spike bait and not a specific antibody (anti-SARS-CoV-2 spike ECLIA assay (Elecsys, Roche Diagnostics), performed by Public Health England, Porton Down).

In agreement with other studies (*15, 16*), our test also confirmed in most individuals that IgG1 is the main responsive immunoglobulin. Figure 2A shows that IgG1 correlates well (r=0.84) across a broad range (10 to 100,000 U/ml) with the Elecsys Anti-S assay.

**Figure 2.**
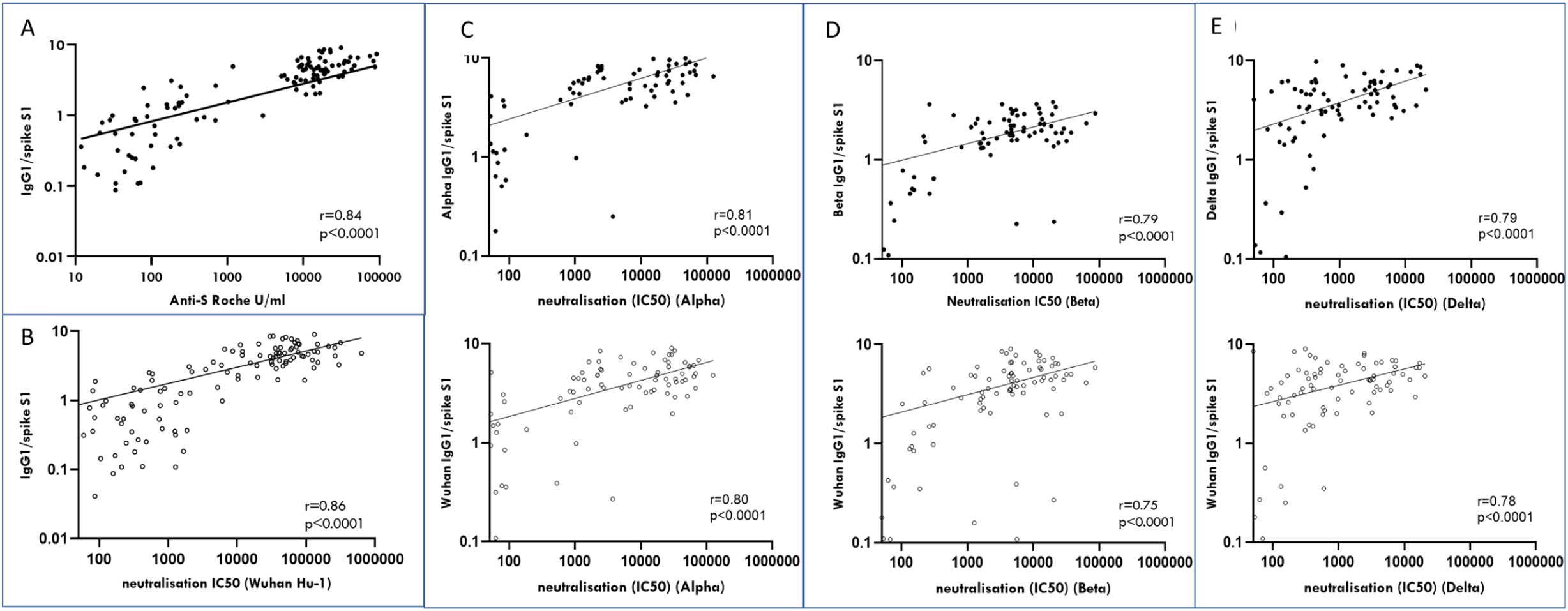
Comparison of LC-MS/MS IgG1 levels with other serology method and live viral neutralisation. (A) Roche Elecsys Anti-S assay vs IgG1 LC-MS/MS (B) Wuhan hu-1 neutralising antibodies vs Wuhan S1 IgG1 LC-MS/MS (C) Alpha neutralising antibodies vs alpha and Wuhan S1 IgG1 LC-MS/MS (D) Beta neutralising antibodies vs Beta and Wuhan S1 IgG1 LC-MS/MS (E) Delta neutralising antibodies vs delta and Wuhan S1 IgG1 LC-MS/MS. Significance determined by spearman correlation. n= 141 in total for n= 24 infection naïve group, n=23 previous infection group at pre, first and second vaccination time points.

The ability to measure all the components of the immunocomplex, demonstrated that IgG1 showed the strongest correlation with live virus neutralisation data (NAbs) (supplementary figure S3) followed by C1q. Figure 2B-E shows comparison of IgG1 against each S1 variant bait correlates well with corresponding NAbs (r=0.79-0.86). Comparison using only IgG1 Wuhan hu-1 S1 against neutralisation data for each variant shows only slightly less correlation against Alpha, Beta and Delta VoC (r=0.75-0.80). This confirms our high throughput assay demonstrated excellent correlation with conventional neutralising cell-based assays.

### Determining the immuno-response and protection to both Wuhan Hu-1 and other variants of concern

Our results for IgG1 (figure 3A) using the LC-MS/MS assay are in line with our published findings that 2 exposures, either via a two dose vaccination protocol, or natural infection and one dose vaccination, have the same effect whereby the second antigen exposure increases anti-Wuhan spike antibody levels on average 3-10 fold (*8, 11, 13*). In concordance with the gold standard S1-RBD binding and authentic live virus neutralisation assays (*8, 13*), we found no increase in anti-Wuhan Hu-1 S1 IgG1 responses from the second vaccine dose in previously infected HCW. This suggesting an antibody ceiling is achieved at third antigen exposure.

**Figure 3.**
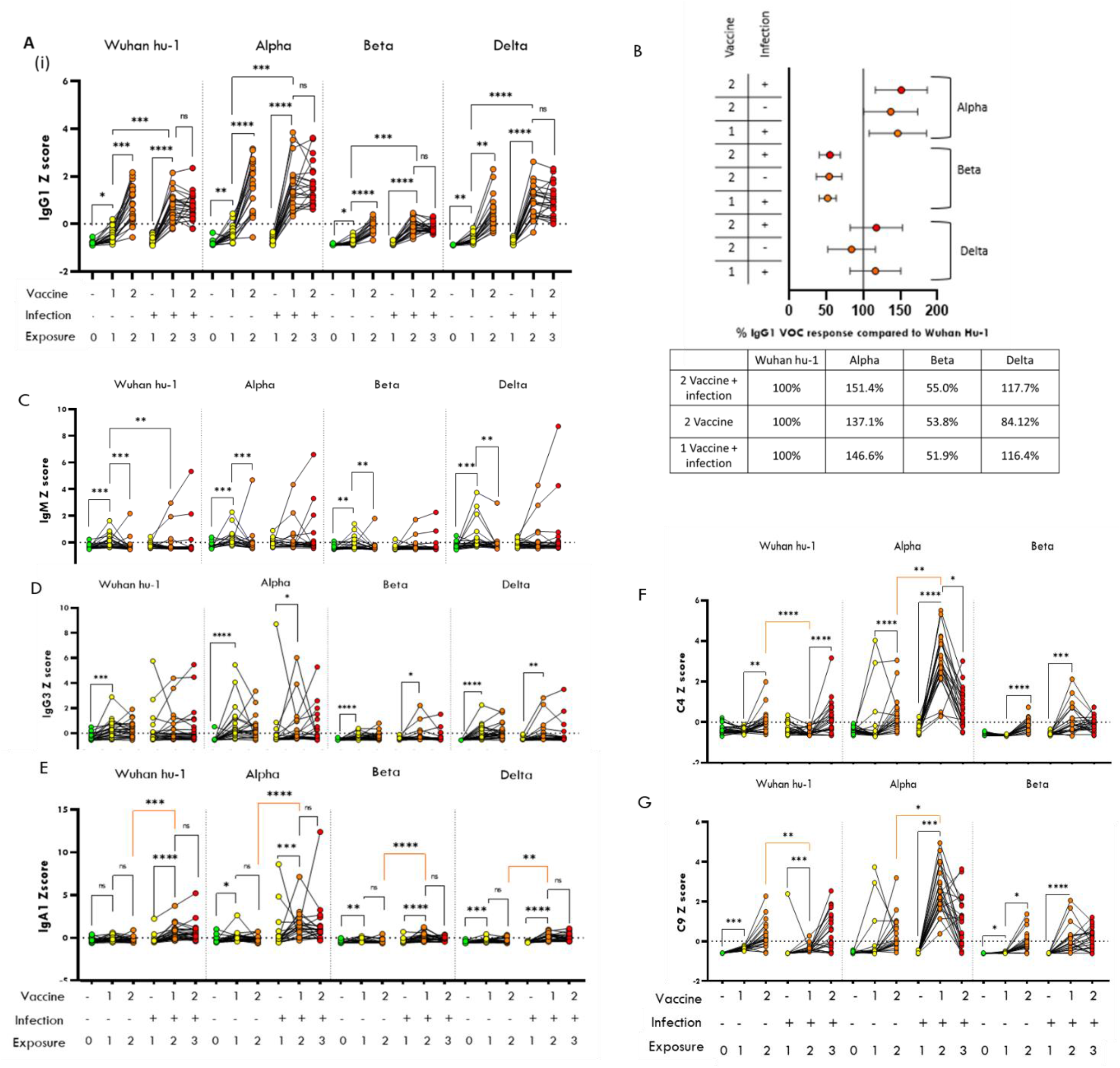
Application of Immunocomplex assay to vaccinated participant cohort. **(A)** Shows the comparison of IgG1 levels binding to each VoC spike according to vaccine status in healthcare workers, who have had (+) or not had (-), a prior SARS CoV2 infection. Results are shown for the wild type Wuhan hu-1, Alpha, Beta and Delta spike variants. **(B)** summarises as an average percentage the IgG1 response to the VoC compared to Wuhan Hu-1. Comparison of other proteins of immunocomplex according to vaccine status for **(C)** IgM **(D)** IgG3 **(E)** IgA1 **(F)** complement C4 and **(G)** complement C9. Vaccination groups coloured according to exposure status. **Green** no exposure, **yellow** 1^st^ exposure (X1), **orange** 2nd exposure (X2) and **red** 3^rd^ exposure(X3). Data normalised for VoC comparison. Significance determined by non-parametric ANOVA and spearman correlation. Coloured significance bars indicate change due to pre-infection. n= 24 infection naïve group, n=23 previous infection group

To determine an individual’s antibody reactivity against existing and new VoC, the S1 protein from each VoC (Alpha, Beta, Delta) were bound in three separate wells to compare a patient’s serum antibody binding capability and immune response. IgG1 values were then presented as a percentage of binding capability to the original Wuhan Hu-1 strain and vaccine target, to give a patient a measure of IgG1 reactivity against each VoC (figure 3B). We observe more than a 37-51% greater response against the Alpha S1 compared to the Wuhan Hu-1 as described previously and which is slightly enhanced by previous natural infection (*2*). IgG1 response to Delta S1 is similar to Wuhan-Hu-1 (-/+17%) and is also enhanced by previous infection (figure 3B). IgG1 response to Beta S1 only elicits a 52-55% response relative to the protection against the Wuhan Hu-1 VoC which is likely due to combined effects of the K417N and N501Y mutations in the Beta spike (*17*). Changes in IgM (figure 3C) are only observed in the infection naïve group for all variants where the levels increase at first vaccination, but levels are lower at second vaccination confirming IgM only changes in response to first exposure. No significant group changes related to exposure or infection were observed for IgG2 or IgG4 in most individuals. However, some individuals had notable and significantly higher levels of IgG2 and IgG4 (supplementary figure S1) and indicate a more personalised medicine approach may be applicable for some individuals. IgG3 is always observed albeit at lower levels and appears to increase at first exposure in the vaccine naïve group and unlike IgM the levels appear to stay elevated with further exposure. The pre-infection group show a small significant increase at first vaccination only for the VoC (figure 3D). Of note are differences in the levels of IgA1 (figure 3E). No change is seen in the infection naïve group against the Wuhan Hu-1 bait but an increase is observed for the pre-infection group at first vaccination. However, for the VoC baits after first vaccination we observed a small increase of IgA1 in the infection naive group. The response in the group who had a previous infection was greater. When comparing double exposure groups (2 dose vaccination infection naïve vs 1 dose vaccination with prior infection), we clearly see a greater IgA1 response (p<0.01) in those individuals who had a natural infection (figure 3E).

### Understanding antibody protection beyond immunoglobulins: Determining and quantitating the complement response against Wuhan Hu-1 and Variants of Concern

Current tests only determine IgG or total response. Using our assay we are able to also determine the significant and important contribution from the complement system. Complement C4 levels demonstrated an average 2-6 fold significant increase against the native Wuhan hu-1. This was observed only at the second vaccination stage in both groups indicating a response to vaccine but interestingly not from exposure to SARS-CoV-2 in natural infection (figure 3F). Complement C9 also appears to significantly increase with vaccination first dose against Wuhan-Hu1 by an average 3.8-5.2 fold (figure 3G). However, the C4 and C9 response to the Alpha and Beta VoC, respectively, were observed to be markedly different than that to the native Wuhan-Hu1. The response to the Alpha shows that both complement C4 and C9 increase greatly in the pre-infection group after first vaccination (9.6-fold and 93-fold, respectively). However, this response is not as great after a second vaccination with a lower 3.8-fold and 46-fold increase, for C4 and C9 respectively compared to the pre-vaccine group. A similar but lower level response pattern is observed for Beta. This indicates pre-infection elicits greater C4 and C9 against VoC binding after a second antigen exposure. Complement C1q binding to all variants increases after exposure similar to that of IgG1 (figure 4A) but the levels observed are lower against the Beta and Delta. Whilst it is not significant, we did observe less C1q binding at 3^rd^ exposure where binding increased 42-fold after first vaccination but only a 31-fold increase was observed at second vaccination in the pre-infected group. The similar pattern of C1q binding to IgG1 is likely due to the direct interaction of C1q with the fc region of IgG, when compared we observe a significant correlation (r>0.8) between IgG1 and C1q for all variants. However, when comparing correlation between variants, a reduction in the ratio of IgG1 to C1q against the Delta VoC and to a lesser extent against the Beta VoC were observed (figure 4B). This suggests potentially a reduced interaction of C1q with IgG1 against Beta and Delta VoC. Considering the relationship of IgG1 to NAbs (r>0.79 figure 2) we also looked at the C1q relationship to NAb levels for each variant (figure 4C). Unlike with the IgG1 levels, we did not observe any significant differences in the gradient/ratio of C1q to NAbs when compared to the other variants. This indicates a better association of C1q to NAbs than IgG1 for the Delta VoC.

**Figure 4.**
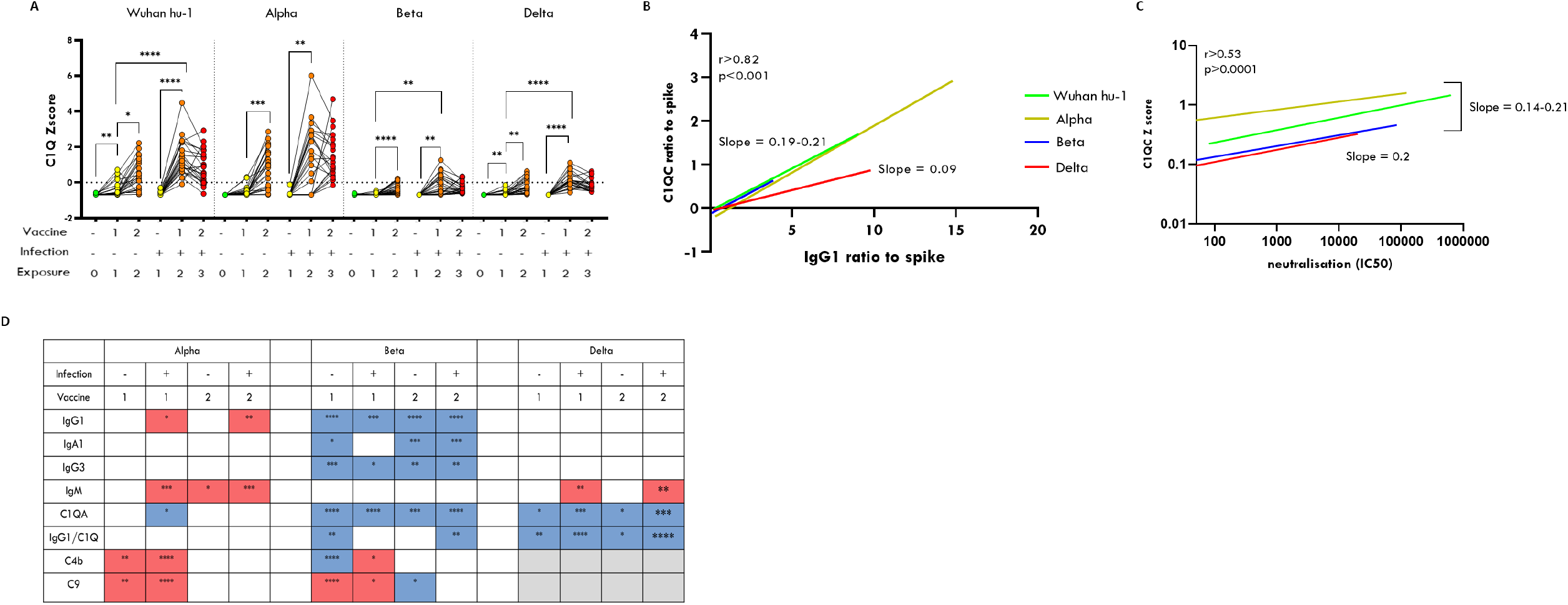
C1q response to variant S1 protein. **(A)** Shows the comparison of C1Q levels binding to each VoC spike according to vaccine status in healthcare workers, who have had (+) (n=23) or not had (-) (n=24), a prior SARS CoV2 infection. Results are shown for the wild type Wuhan hu-1, Alpha, Beta and Delta spike variants. Linear regression based on correlation showing changes in the slope/ratio for Delta for (**B)** C1Q vs IgG1 and no change in slope/ratio for Delta for **(C)** C1Q vs nAbs (log scale) analysis performed on all 4 vaccination groups combined n=94. **(D)** shows a summary of significant changes of the immunocomplex due to VoC compared to Wuhan-Hu-1 response. Blue decreased and red increased. Vaccination groups coloured according to exposure status. **Green** no exposure, **yellow** 1^st^ exposure, **orange** 2nd exposure and **red** 3^rd^ exposure. Significance determined by non-parametric ANOVA and spearman correlation.

To demonstrate this further, when we combine the NAb data for all variants and then compare with IgG1, and also C1q, we can observe that C1q has an overall stronger correlation (r=0.62 vs 0.71) with NAb than IgG1 (supplementary figure S4A-B). If we look at the data as a ratio of C1q to IgG1 across the vaccine groups, we see this ratio is increased with previous exposure to infection (p<0.01 all variants) but this significantly declines at third exposure (p<0.001 all variants) (supplementary figure S4C). When comparing across the variants, we see that the Beta and Delta groups show a significant reduced ratio compared to Wuhan Hu-1, with the exception of the second exposure groups where the ratios are normal against the Beta VoC (supplementary figure S4C). The significance of this observation is unknown and requires further investigation but could relate to a change in the antibody function or maturation that’s affected by prior infection and variant type.

The VoC response compared to Wuhan-hu1 is summarised in figure 3H showing an overall reduced response to the Beta VoC although with a greater C9 response at first vaccination. A greater IgM response against Alpha and Delta is detected in the pre-infected groups and is likely due to a first exposure to a variant. These changes likely relate to contribution of non-neutralising antibodies that use complement mediated lysis which can be driven by IgM (*17*).

## Discussion

Standard SARS-CoV-2 serological assays are becoming less effective as a representation of a ‘correlate of protection’ against variants (*2*). This is due to the lack of specificity of many tests which may either measure a total response to bait antigen such as the Roche Elecsys assay or total IgG. A measure of total response of all proteins with affinity for the spike protein will include all other components of the immunocomplex, such as other class immunoglobulins and complement. However, these conventional assays do not tell us how much these immune proteins contribute individually. Whilst the total response has been adequate for monitoring response to the original Wuhan hu-1, it is no longer adequate against VoC and in populations with an evolving immune response to multiple exposures (*2*).

Our multiplexed approach is capable of looking at all immune reactive proteins individually and demonstrates that other classes of immune proteins, such as IgA and the complement system, also significantly contribute to an individual’s overall immune reactivity against VoC, all of which would be missed by conventional testing.

Just looking specifically at IgG1 if we use the Wuhan hu-1 response as a baseline we see a slight significant increase of IgG1 against the, Alpha VoC (figure 2B and 4D) which could be due to a more open conformation of the RBD domain in the Alpha spike (*18*). However we observe no change for the Delta VoC. In addition and confirming previous findings, pre-infection has been shown to significantly boost antibody response of the first dose of the vaccination (*8, 19, 20*). However, our data indicates that a double vaccine dose in the infection naïve, still does not achieve mean values of those who had pre-infection. Thus the combination of infection plus vaccination may provide a potentially higher immunity versus that of a conventional 2 dose vaccination protocol. This effect is more noticeable for Delta exposure (figure 2B) and previous reports have shown that prior infection does reduce the rate of Delta breakthrough infections (*21*).

However contrary to our observation of greater or unaltered IgG1 it has been shown that NAbs are lower against VoC (*2*). This indicates IgG1 is less of a reflection of NAb ability against the VoC. This is likely due to a greater contribution of non-neutralising antibodies against the VoC that can’t be differentiated by IgG1 measurement.

The evaluation of bound antibodies and complement fixation beyond just IgG1 in our assay, revealed a surprisingly large degree of heterogeneity in response to exposure with some individuals having completely altered immunoglobulin profiles (supplementary figure S1). The relevance of some patients having a lower IgG1 but higher IgG2 and IgG4 response is unknown, but this observation would be overlooked using conventional testing. This could also have relevance in future studies of inter-individual and longitudinal responses to continued exposure of SARS-CoV-2 and vaccines. This ability to define an individual’s immunoglobulin profile and determine the variability of an individual’s immunological make up, is both intriguing and potentially very important, particularly in different clinical scenarios such as disease severity and mortality (*16, 22, 23*).

Unlike other assays available, our platform also measures the immunocomplex binding from the lesser well understood but very important complement system. Complement activation can contribute to anti-viral defence by the classical or lectin pathway leading to neutralisation by viral opsonisation and lysis (*24–27*). Studies on parainfluenza virus show that complement mediated neutralisation is likely to be a mechanism of non-neutralising antibody action driven by IgM (*24*). In our data, we observe a greater IgM response against Alpha and Delta, which is also accompanied by greater C4 and C9 binding (Figure 4D). This would indicate a greater proportion of non-neutralising antibodies as also determined by the lower concordance of IgG1 with VoC NAbs that we describe earlier (figure 2).

One of the more interesting findings in the analyses of the immunocomplex was that of the role of C1q. C1q directly interacts with the fc portion of immunoglobulins and is required for initiation of the complement cascade. In our analysis C1q binding appears to behave independently from the other complement components C4 and C9 (figure 4) and showing a reduction in binding against the Beta and Delta VoC (figure 4A) this could be due to C4 and C9 binding to virions to opsonise the virus as opposed to C1q which may just be bound to antibody. Interestingly, for the Delta VoC, C1q was reduced relative to IgG1 but not relative to NAbs. This indicates that C1q could be a better surrogate indicator of NAb protection against SARS-CoV2 variants than IgG1. Previous work has shown that heat-inactivation, which would inactivate complement appears not to affect IgG reactivity to the RBD domain (*28*), but may affect its neutralisation capability (*29*). This confirms that complement whilst is not essential may have a role in neutralisation. Further weight to this observation comes from Melhop *et al*, who showed that C1q increases the potency of antibodies against West Nile virus by modulating the stoichiometric requirements for neutralisation (*30*). Therefore, it is possible that complement could contribute potentially to protection against VoC by C1q augmenting neutralising antibodies and non-neutralising antibodies using the classical complement pathway for viral opsonisation. If indeed C1q binding is more specific to NAbs then the altered ratio between C1q to IgG1 for the Delta VoC, which we observed in this study again could be explained by a greater proportion of non-neutralising IgG1 antibodies present. This finding of the C1q association with NAbs merits further investigation to confirm if the IgG1 to C1q relationship could be an indirect way of determining neutralising and non-neutralising antibody responses to VoC.

In this study, our data shows only a snapshot of the antibody response approximately 3 weeks after vaccination in non-hospitalised healthcare workers. Further studies to characterise the effect over time, infection from different variants and infection severity, and how the ‘immunocomplex signature’ changes with age, would give us a greater understanding of the evolving antibody mediated immune response to SARS-CoV-2. Our findings also uncover an area of antibody mediated immunity that is little understood and complement function is far more complex than what we can relay in our study. One of the limitations with our assay is that we are not able to determine functional complement activation, although precise quantitative detection may be able to funnel further investigation. Our approach using multiplex LC-MS/MS could provide a valuable platform to better enable research in this area. Whilst the LC-MS/MS multiplex assay is a research standard assay, it was designed so it can be easily translated for use in a clinical laboratory setting (*7*). Simply, by changing the bait to a VoC spike protein, our test can be easily modified to determine antibody mediated immune response for current and emerging VoC. The information obtained will allow us to understand in greater detail an individual’s antibody protection or be used in vaccine design. Furthermore this ‘Bait, Capture and Mass Spec’ approach, can also have applications beyond SAR-CoV-2 for other infectious diseases, or even immune response to novel treatments and autoimmunity.

## Methods

### Ethics statement

Human sera were obtained from the COVIDsortium Healthcare Workers bioresource [(*8–10, 12, 13*)] which is approved by the ethical committee of UK National Research Ethics Service (20/SC/0149) and registered on ClinicalTrials.gov (NCT04318314). The study conformed to the principles of the Helsinki Declaration, and all subjects gave written informed consent.

### COVIDsortium Healthcare Worker Participants

SARS-CoV-2 infection (by the Wuhan Hu-1 strain) of study participants was determined by baseline and weekly nasal RNA stabilizing swabs and Roche cobas® SARS-CoV-2 reverse transcriptase polymerase chain reaction (RT-PCR) test as well as baseline and weekly serology using the spike protein S1 IgG EUROIMMUN enzyme-linked immunosorbent assay (ELISA) and nucleocapsid ROCHE Elecsys electrochemiluminescence immunoassay (ECLIA). Antibody ratios >1.1 were considered test positive for the EUROIMMUN SARS-CoV-2 ELISA and >1 was considered test positive for the ROCHE Elecsys anti-SARS-CoV-2 ECLIA following Public Health England evaluation (*8–11, 31*)

The previously reported^13,14^ cross-sectional, case-controlled vaccine sub-study (n=51) collected samples at a mean/median timepoint of 22d and 20d after administration of the first and second dose of the mRNA vaccine, BNT162b2. This vaccine sub-study recruited HCW previously enrolled in the 16-18 week sub-study(*11*). This included 25 HCW (mean age 44yr, 60% male) with previous laboratory defined evidence of WT SARS-CoV-2 infection and twenty-six HCW (mean age 41y, 54% male) with no laboratory evidence of SARS-CoV-2 infection throughout the initial 16-week longitudinal follow up. Neutralising antibody and RBD ELISA data obtained by ROCHE Elecsys anti-SARS-CoV-2 ECLIA following Public Health England (PHE) has been previously published (*8–12*).

### SARS2-CoV-2 Immunocomplex assay

#### Bait capture

Ninety six well microtitre plates (Waters Corp) were coated with either Wuhan Hu-1 SARS-CoV-2 spike protein (S1) (Genscript Z03501), Alpha VOC B.1.1.7 SARS-CoV-2 (2019-nCoV) Spike S1 (HV69-70 deletion, N501Y, D614G)-His Recombinant Protein (Sino biological 40591-V08H7), Beta VOC B.1.1351 SARS-CoV-2 Spike protein (S1, E484K, K417N, N501Y, His Tag (Genscript Z03531-1) or Delta VOC B.1.617.2 (T19R, G142D, E156G, 157-158 deletion, L452R, T478K, D614G, P681R) Protein (His Tag) (Sino biological 40591-V08H19). S1 protein was diluted to 50 µg/ml in PBS and 10 µl added to the bottom of each well. Wells were topped with 140 µl of Carbonate/Bicarbonate buffer 100 mM at pH 9.6 and then incubated for 12-16 hours at 4 °C. All further incubations were performed at room temperature (RT) unless stated otherwise. Supernatant was carefully tipped from the wells. Wells were washed with 200 µl of PBS and then incubated for 1 hr with 200 µl of blocking solution consisting 1mg/ml of horse myoglobin (Sigma UK) in PBS followed by 3 washes with PBS. Plate wells were stored with PBS and kept at 4 °C until used.

Serum samples were diluted 1:10 in 0.05 mg/ml horse myoglobin (Sigma UK) in PBS and added to S1 protein coated wells for 1hr at 37°C. Sample was carefully removed and wells were washed once with 200 µl 0.05 % Tween 20 in PBS and then 3 times with PBS. For saliva 75 l of neat saliva was diluted 1:1 in 0.05 mg/ml horse myoglobin in PBS and added to baited wells.

Dried blood spots: 6 mm DBS spots were punched into a 2 ml micro tube and extracted using 175 µl of 0.05mg/ml Horse myoglobin solution in PBS for 1 hour on the shaker. Samples were centrifuged at max rpm on benchtop centrifuge for 10 min. An aliquot 150 µl per reaction was added to the baited well for 1hr.

Dried Saliva spot (lollipop) – 6 mm saliva spots were punched into a 2 ml micro tube and extracted using 250 µl of 0.05mg/ml Horse myoglobin solution in PBS for 1 hour on the shaker. Centrifuge at max rpm on benchtop centrifuge for 10 min. An aliquot of 150 µl per reaction was added to the baited well for 1hr.

#### Immunocomplex protein digestion

Seventy microliters of 0.5 % Sodium deoxycholate in 50 mM Ammonium Bicarbonate buffer was added to each well followed by 3 µl of DTT solution (DL-Dithiothreitol (DTT) – 162 mM in 0.5 % sodium deoxycholate/ 50 mM Ammonium bicarbonate buffer). Plates were capped and incubated at 85°C for 15 min with shaking (750 rpm). The plate was left to cool to room temperature before 6 µl of 162 mM Iodoacetamide (IAA) in 0.5 % Sodium deoxycholate/ 50 mM Ammonium bicarbonate buffer was added. Plates were capped and brief shaken and incubated at room temperature for 30 min. Five microliters of trypsin (Sigma) (1 mg/ml in 50 mM Acetic Acid) was added and incubated at 45°C for 30 mins. Digestion was halted by addition of 5 µl of 6% TFA and mixed well. Plates were centrifuged for 20 min at 4000 g at 10°C. Fifty microliters are aliquoted into a fresh plate and analysed by LC-MS/MS.

#### Targeted LC-MS/MS analysis

Digested samples were injected onto a Waters 50 mm UPLC Premier ® C18 1.7 µm, 2.1 × 50 mm column operating at 45 °C, for chromatographic separation. Mobile phase A consisted of: 0.1% formic acid in water and B: 0.1% formic acid in ACN, pumped at a flow rate of 0.3 mL min-1. The starting conditions of 5% B were kept static for 0.1 minutes, before initialising the linear gradient to elute and separate peptides over 7.7 minutes to 40% B. B was linearly increased to 80% over 0.2 minutes and held for 1 minute to wash the column before returning to the initial conditions followed by equilibration for 1 minute prior to the subsequent injection. The LC system was coupled to a Waters Xevo-TQ-S triple quadrupole mass spectrometer for multiple reaction monitoring (MRM) detection in positive electrospray ionisation mode. The capillary voltage was set to 2.8 kV, the source temperature to 150 °C, the desolvation temperature to 600 °C, the cone gas and desolvation gas flows to 150 and 800 L hour-1 respectively. The collision gas consisted of nitrogen and was set to 0.15 mL min-1. The nebuliser operated at 7 bar. The cone energy was set to 35 V and the collision energies varied depending on the optimal settings for each peptide. Transition information of each peptide is given in supplementary file 1.

#### Authentic Virus Neutralisation Assay

SARS-CoV-2 microneutralisation assays were previously reported(*2, 8*). VeroE6 cells were seeded in 96-well plates 24h prior to infection. Duplicate titrations of heat-inactivated participant sera were incubated with 3×10^4^ FFU SARS-CoV-2 virus (TCID100) at 37°C, 1h. Serum/virus preparations were added to cells and incubated for 72h. Surviving cells were fixed in formaldehyde and stained with 0.1% (wt/vol) crystal violet solution (crystal violet was resolubilised in 1% (wt/vol) sodium dodecyl sulphate solution). Absorbance readings were taken at 570nm using a CLARIOStar Plate Reader (BMG Labtech). Negative controls of pooled pre-pandemic sera (collected prior to 2008), and pooled serum from neutralisation positive SARS-CoV-2 convalescent individuals were spaced across the plates. Absorbance for each well was standardised against technical positive (virus control) and negative (cells only) controls on each plate to determine percentage neutralisation values. IC50s were determined from neutralisation curves. All authentic SARS-CoV-2 propagation and microneutralisation assays were performed in a containment level 3 facility.

#### Data Analysis

Raw LC-MS/MS data was analysed using Skyline open source software (https://skyline.ms/project/home/software/Skyline/begin.view). Peptide identifications were determined from prior analysis of digested serum and immunoglobulin standards (Invitrogen) by a minimum of 6 transitions and matched to *in-silico* spectral library (Prosit) for additional confirmation. Two optimal transitions were used for final MRM analysis. Peptide abundance data were normalised to S1 peptide FASVYAWNR which was present in all variants. For comparison analysis between variant assays ratio values were normalised by Z-Score. Exported data were analysed using Microsoft Excel and Graphpad Prism v9.

Linearity response of IgG upto 250 g/ml (r>0.99) was confirmed using a calibration curve using IgG protein standard (Sigma, UK) the maximum observed sample IgG1 value was 157.23 ug/ml well within linear range. LOD and LOQ values (supplementary table 1) were determined by proportion of each IgG subclass of standard. High QC and LQC serum was obtained from vaccinated volunteers. Immunoglobulin CVs were below 30% for all variants apart from low levels for IgG3 and 4. Complement proteins were only detectable in the HQC and showed freeze thaw instability of C4-C9. Therefore complement data for the later emerging Delta variant is not shown.

#### Statistics

For comparison of vaccination groups nonparametric ANOVA (Kruskal Wallis) were used to determine significance. For correlation analyses nonparametric spearman test was used to determine significance and r value.

## Supporting information

Members of covidsortium

## Data Availability

All data produced in the present study are available upon reasonable request to the authors

## Supplementary Material

Supplementary information: List of Covidsortium members

Supplementary figure S1. Multivariate analysis of the immunocomplex response to SARS-CoV2 infection or vaccine.

Supplementary figure S2. Proof of principle of application of assay to other tissues.

Supplementary Figure S3. Spearman correlation matrix of components of the immunocomplex and corresponding neutralising antibodies.

Supplementary Figure S4. C1q relationship to IgG1 and neutralising antibodies.

## Acknowledgments

The views expressed are those of the author(s) and not necessarily those of the NHS, the NIHR, UKRI or the Department of Health. We wish to thank Drs Mike Morris and Don Cooper at Waters for their advice and consumables support. We would also like to thank Dr Rachel Carling for her advice on the clinical translation adaption of the assay and to the members of the Covidsortium (see supplementary information). Funding: This work is (partly) funded by the NIHR GOSH BRC, TMSRG UCL and The Peto Foundation. RJB, DMA and AMK are supported by UKRI MR/W020610/1.

## Author information

Conceptualisation: KM

Methodology: KG, WH

Formal analysis: ID, WH

Investigation: ID, JS, JMG, CP, RJB, TT, CM, GC, MF, AS

Resources: KG, RJB, TT, CM, JCM, GC, MF, TB

Data Curation: RJB, TT, CM, GC, MF

Writing – Original Draft: WH

Writing - Review & Editing: MN, AMK, RJB, DMA, MKM

Supervision: WH, KM, JCM

Funding acquisition: JCM, KM

## Competing interests

The authors have submitted an intellectual property claim for using the technology for clinical applications.

## Data availability

The dataset supporting the findings of this study are available as an Extended Data file. MRM assay source data are available at https://panoramaweb.org/SARCOV2immunocomplex.url

## Notes

### Competing Interest Statement

The authors have submitted an intellectual property claim for using the platform for clinical applications.

### Author Declarations

Human sera were obtained from the COVIDsortium Healthcare Workers bioresource which is approved by the ethical committee of UK National Research Ethics Service (20/SC/0149) and registered on ClinicalTrials.gov (NCT04318314). The study conformed to the principles of the Helsinki Declaration, and all subjects gave written informed consent.

## References

1. C. Gaebler et al., Evolution of antibody immunity to SARS-CoV-2. Nature 591, 639–644 (2021).

2. C. J. Reynolds et al., Heterologous infection and vaccination shapes immunity against SARS-CoV-2 variants. Science, eabm0811 (2021).

3. J. Abbasi, The Flawed Science of Antibody Testing for SARS-CoV-2 Immunity. JAMA 326, 1781–1782 (2021).

4. J. Nie et al., Quantification of SARS-CoV-2 neutralizing antibody by a pseudotyped virus-based assay. Nat Protoc 15, 3699–3715 (2020).

5. H. Q. Yu et al., Distinct features of SARS-CoV-2-specific IgA response in COVID-19 patients. Eur Respir J, (2020).

6. B. Ju et al., Human neutralizing antibodies elicited by SARS-CoV-2 infection. Nature 584, 115–119 (2020).

7. N. P. M. Smit et al., The Time Has Come for Quantitative Protein Mass Spectrometry Tests That Target Unmet Clinical Needs. J Am Soc Mass Spectrom 32, 636–647 (2021).

8. C. J. Reynolds et al., Prior SARS-CoV-2 infection rescues B and T cell responses to variants after first vaccine dose. Science, (2021).

9. T. A. Treibel et al., COVID-19: PCR screening of asymptomatic health-care workers at London hospital. Lancet 395, 1608–1610 (2020).

10. C. Manisty et al., Antibody response to first BNT162b2 dose in previously SARS-CoV-2-infected individuals. Lancet 397, 1057–1058 (2021).

11. C. Manisty et al., Time series analysis and mechanistic modelling of heterogeneity and sero-reversion in antibody responses to mild SARSCoV-2 infection. EBioMedicine 65, 103259 (2021).

12. C. J. Reynolds et al., Discordant neutralizing antibody and T cell responses in asymptomatic and mild SARS-CoV-2 infection. Sci Immunol 5, (2020).

13. e. a. Reynolds, Heterologous B.1.1.7 infection and two-dose vaccination reduces immunity against other variants. Science Immediate Release, December 2nd, 2021, (2021).

14. M. van der Neut Kolfschoten et al., Anti-inflammatory activity of human IgG4 antibodies by dynamic Fab arm exchange. Science 317, 1554–1557 (2007).

15. M. Dogan et al., SARS-CoV-2 specific antibody and neutralization assays reveal the wide range of the humoral immune response to virus. Commun Biol 4, 129 (2021).

16. H. P. Patil et al., Antibody (IgA, IgG, and IgG Subtype) Responses to SARS-CoV-2 in Severe and Nonsevere COVID-19 Patients. Viral Immunol 34, 201–209 (2021).

17. D. Zhou et al., Evidence of escape of SARS-CoV-2 variant B.1.351 from natural and vaccine-induced sera. Cell 184, 2348–2361 e2346 (2021).

18. T. J. Yang et al., Effect of SARS-CoV-2 B.1.1.7 mutations on spike protein structure and function. Nat Struct Mol Biol 28, 731–739 (2021).

19. G. Anichini et al., SARS-CoV-2 Antibody Response in Persons with Past Natural Infection. The New England journal of medicine 385, 90–92 (2021).

20. J. E. Ebinger et al., Antibody responses to the BNT162b2 mRNA vaccine in individuals previously infected with SARS-CoV-2. Nature medicine 27, 981–984 (2021).

21. P. Kim, S. M. Gordon, M. M. Sheehan, M. B. Rothberg, Duration of SARS-CoV-2 Natural Immunity and Protection against the Delta Variant: A Retrospective Cohort Study. Clin Infect Dis, (2021).

22. E. Della-Torre et al., Serum IgG4 level predicts COVID-19 related mortality. Eur J Intern Med 93, 107–109 (2021).

23. M. Perez-Toledo et al., SARS-CoV-2-specific IgG1/IgG3 but not IgM in children with Pediatric Inflammatory Multi-System Syndrome. Pediatr Allergy Immunol 32, 1125–1129 (2021).

24. S. Vasantha et al., Interactions of a nonneutralizing IgM antibody and complement in parainfluenza virus neutralization. Virology 167, 433–441 (1988).

25. J. P. Jayasekera, E. A. Moseman, M. C. Carroll, Natural antibody and complement mediate neutralization of influenza virus in the absence of prior immunity. J Virol 81, 3487–3494 (2007).

26. U. Kunnakkadan et al., Complement-Mediated Neutralization of a Potent Neurotropic Human Pathogen, Chandipura Virus, Is Dependent on C1q. J Virol 93, (2019).

27. B. Schiela et al., Active Human Complement Reduces the Zika Virus Load via Formation of the Membrane-Attack Complex. Frontiers in immunology 9, 2177 (2018).

28. F. Amanat et al., A serological assay to detect SARS-CoV-2 seroconversion in humans. medRxiv, (2020).

29. B. Pastorino, F. Touret, M. Gilles, X. de Lamballerie, R. N. Charrel, Heat Inactivation of Different Types of SARS-CoV-2 Samples: What Protocols for Biosafety, Molecular Detection and Serological Diagnostics? Viruses 12, (2020).

30. E. Mehlhop et al., Complement protein C1q reduces the stoichiometric threshold for antibody-mediated neutralization of West Nile virus. Cell Host Microbe 6, 381–391 (2009).

31. Public Health England, COVID-19: laboratory evaluations of serological assays, gov.uk, 16 March 2021 (www.gov.uk/government/publications/covid-19-laboratory-evaluations-ofserological-assays.).

