## Supplementary figures S1-S4 for "Quantitative, multiplexed, targeted proteomics for ascertaining variant specific SARS-CoV-2 antibody response"

Supplementary figure S1. Multivariate analysis of the immunocomplex response to SARS-CoV2 infection or vaccine.

Supplementary figure S2. Proof of principle of application of assay to other tissues.

Supplementary Figure S3. Spearman correlation matrix of components of the immunocomplex and corresponding neutralising antibodies .

Supplementary Figure S4. C1q relationship to IgG1 and neutralising antibodies.

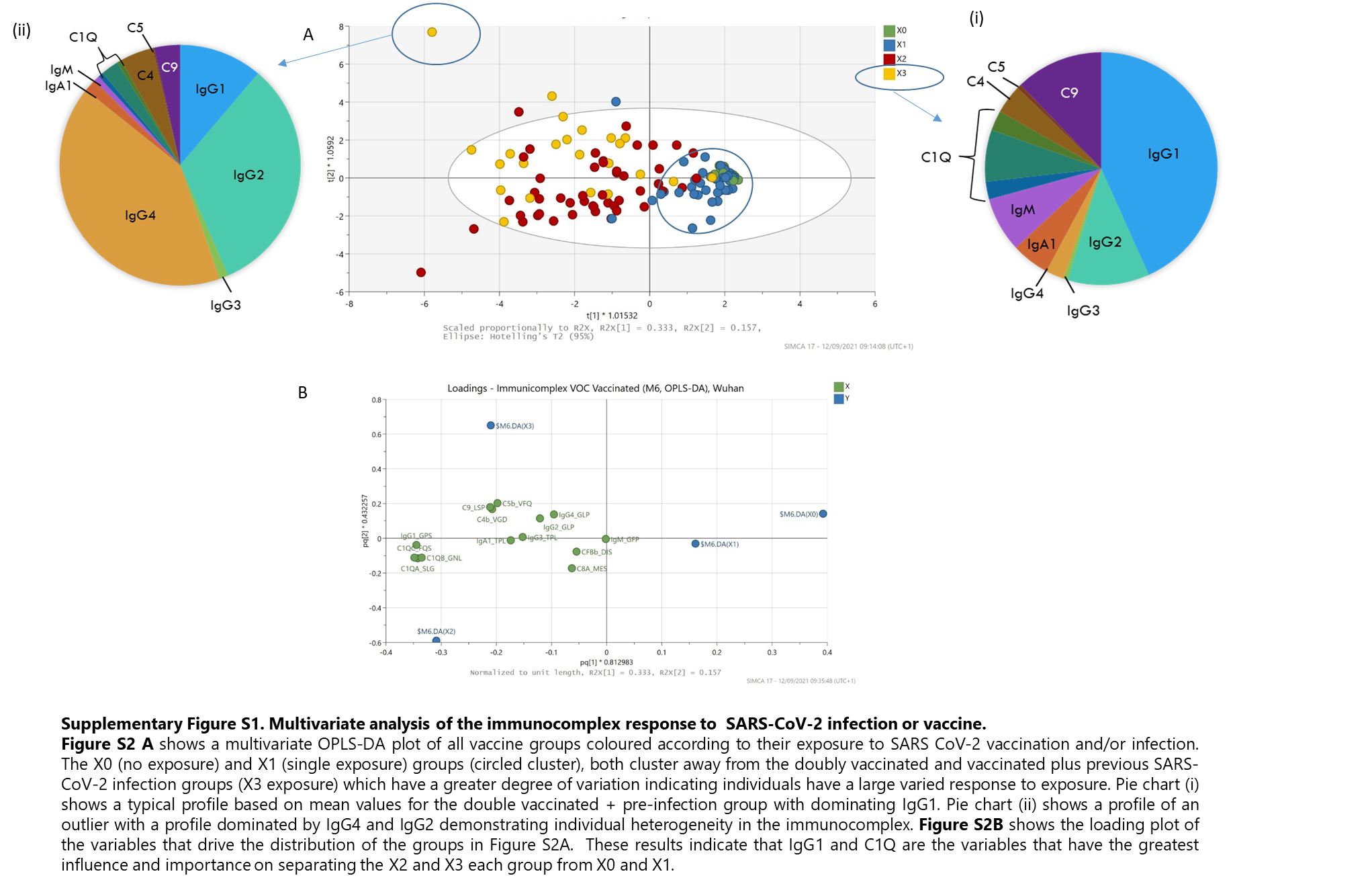

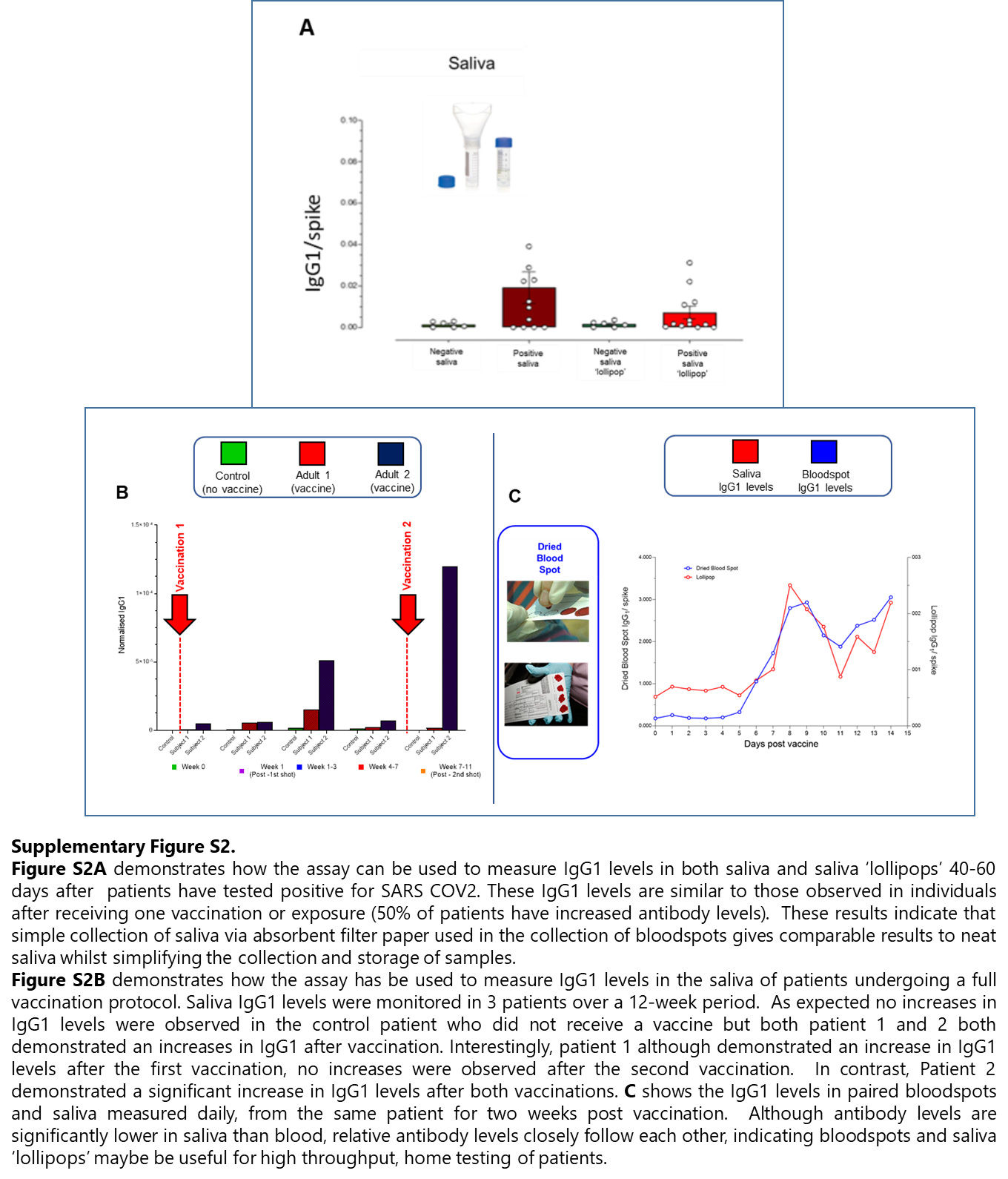

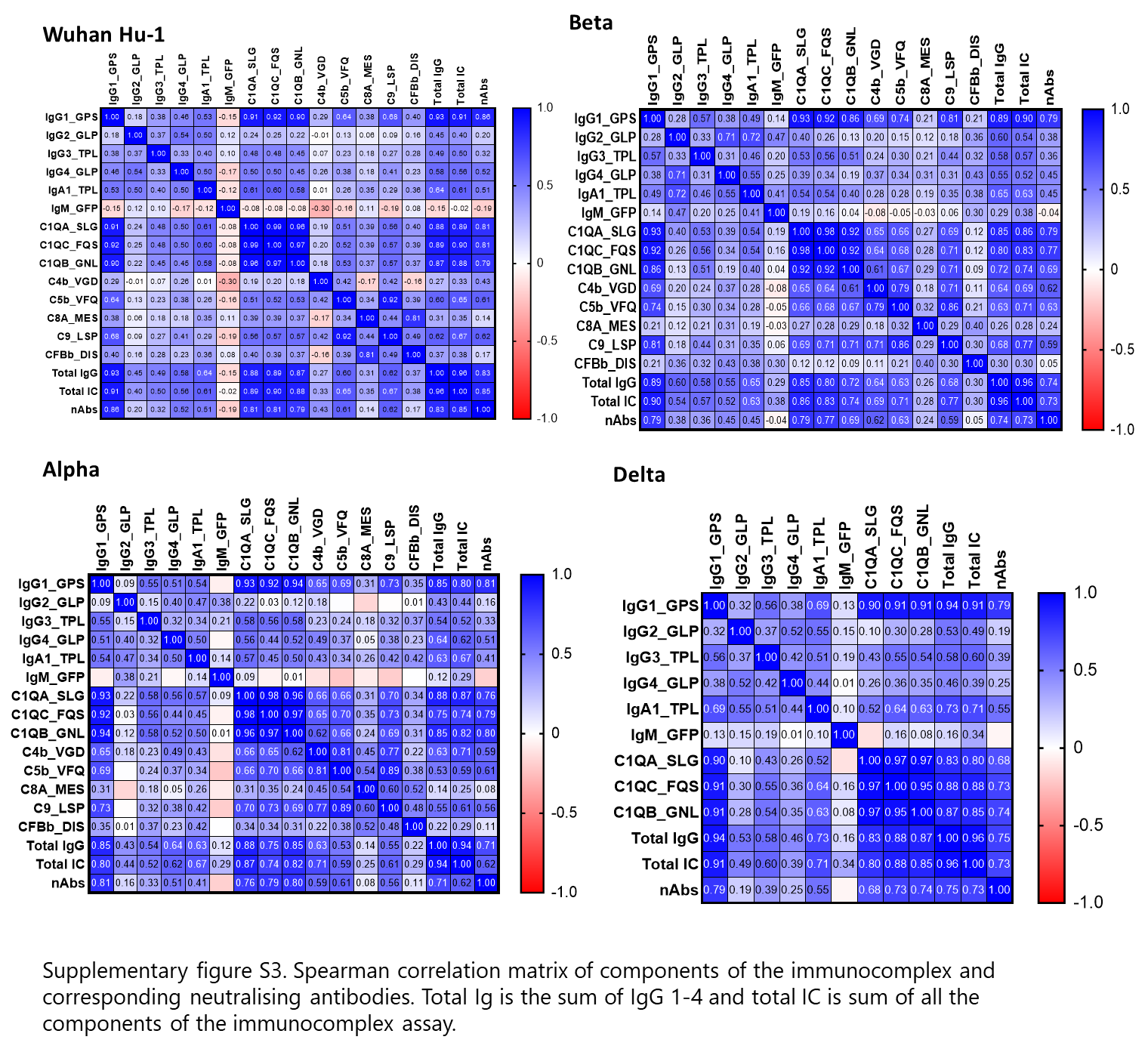

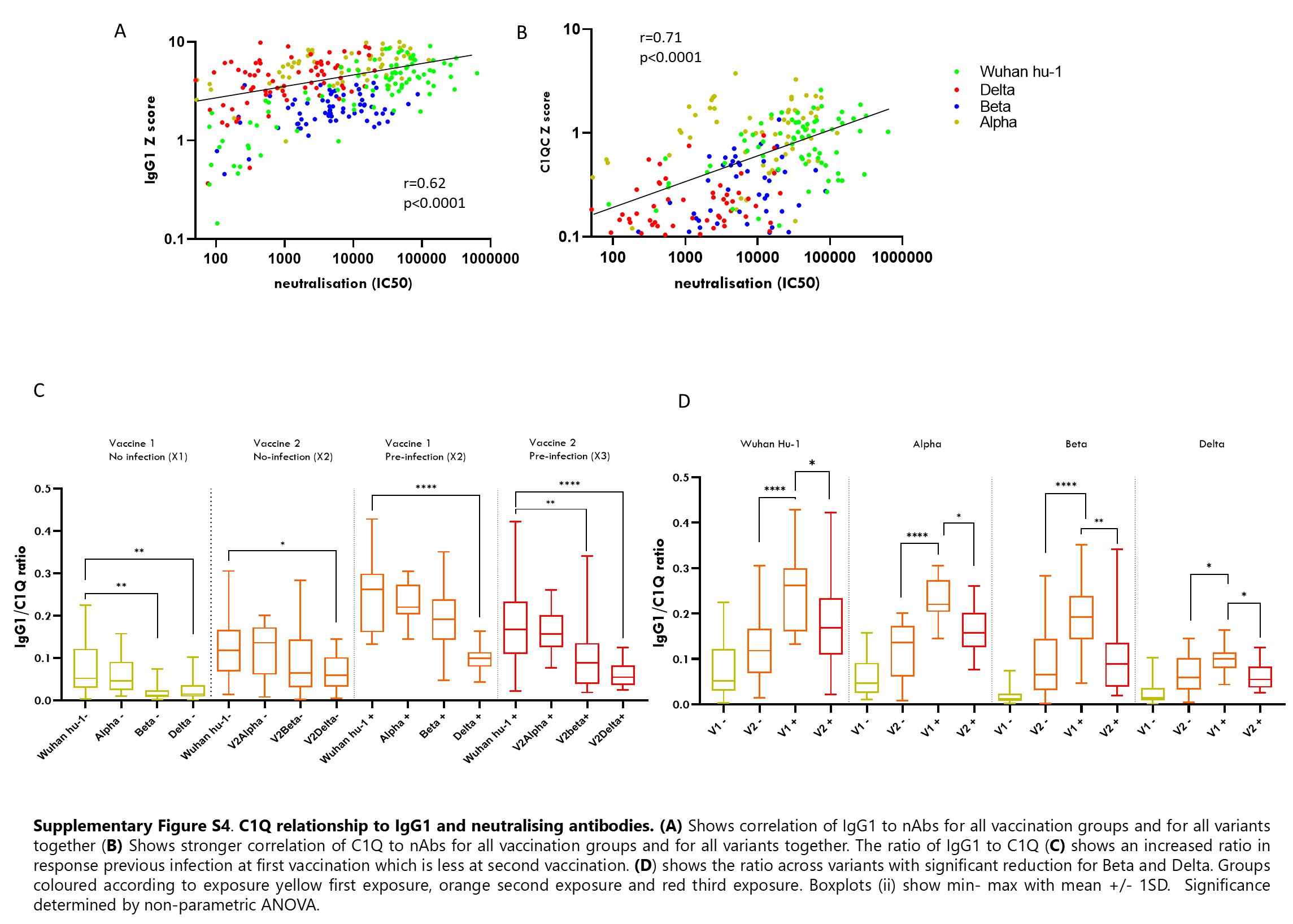
