## Supplementary material for "Quantitative, multiplexed, targeted proteomics for ascertaining variant specific SARS-CoV-2 antibody response": Members of covidsortium

### The members of the UK COVIDsortium investigators are:

Hakam Abbass, Aderonke Abiodun, Mashael Alfarih, Zoe Alldis, Daniel M Altmann, Oliver E Amin, Mervyn Andiapen, Jessica Artico, João B Augusto, Georgina L Baca, Sasha N L. Bailey, Anish N Bhuva, Alex Boulter, Ruth Bowles, Rosemary J Boyton, Olivia V Bracken, Ben O’Brien, Tim Brooks, Natalie Bullock, David K Butler, Gabriella Captur, Olivia Carr, Nicola Champion, Carmen Chan, Aneesh Chandran, Tom Coleman, Jorge Couto de Sousa, Xose Couto-Parada, Eleanor Cross, Teresa Cutino-Moguel, Silvia D’Arcangelo, Rhodri H Davies, Brooke Douglas, Cecilia Di Genova, Keenan Dieobi-Anene, Mariana O Diniz, Anaya Ellis, Karen Feehan, Malcolm Finlay, Marianna Fontana, Nasim Forooghi, Joseph M Gibbons, Derek Gilroy, Matt Hamblin, Gabrielle Harker, Jacqueline Hewson, Wendy Heywood, Lauren M Hickling, Bethany Hicks, Aroon D Hingorani, Lee Howes, Ivie Itua, Victor Jardim, Wing-Yiu Jason Lee, Melaniepetra Jensen, Jessica Jones, Meleri Jones, George Joy, Vikas Kapil, Caoimhe Kelly, Hibba Kurdi, Jonathan Lambourne, Kai-Min Lin, Siyi Liu, Sarah Louth, Mala K Maini, Vineela Mandadapu, Charlotte Manisty, Áine McKnight, Katia Menacho, Celina Mfuko, Kevin Mills, Sebastian Millward, Oliver Mitchelmore, Christopher Moon, James Moon, Diana Muñoz Sandoval, Sam M Murray, Mahdad Noursadeghi, Ashley Otter, Corinna Pade, Susana Palma, Ruth Parker, Kush Patel, Mihaela Pawarova, Steffen E Petersen, Brian Piniera, Franziska P Pieper, Lisa Rannigan, Alicja Rapala, Catherine J Reynolds, Amy Richards, Matthew Robathan, Joshua Rosenheim, Cathy Rowe, Jane Sackville West, Genine Sambile, Nathalie M. Schmidt, Amanda Semper, Andreas Seraphim, Mihaela Simion, Angelique Smit, Michelle Sugimoto, Leo Swadling, Stephen Taylor, Nigel Temperton, Stephen Thomas, George D Thornton, Thomas A Treibel, Art Tucker, Ann Varghese, Jessry Veerapen, Mohit Vijayakumar, Tim Warner, Sophie Welch, Theresa Wodehouse, Lucinda Wynne, and Dan Zahedi
